# Are low ergothioneine levels a risk factor for age-related macular degeneration and other ocular disorders?

**DOI:** 10.64898/2026.02.27.26347162

**Authors:** Irwin K. Cheah, Zachary Fong, Lucrecia Chen, Richard M.Y. Tang, Lei Zhou, Yasuo Yanagi, Ching-Yu Cheng, Xinyi Su, Xueying Li, Kelvin Yi Chong Teo, Chui Ming Gemmy Cheung, Tien-En Tan, Barry Halliwell

**Author notes:** Both authors contributed equally to this work (Irwin K. Cheah, Zachary Fong). **Co-corresponding authors: Prof. Barry Halliwell:**, Address: Centre for Life Science, 28 Medical Drive, #05-01, National University of Singapore, 117456, Singapore., **Dr. Tien-En Tan:**, Address: Singapore National Eye Centre, 11 Third Hospital Avenue, 168751, Singapore., **Prof. Chui Ming Gemmy Cheung:**, Address: Singapore National Eye Centre, 11 Third Hospital Avenue, 168751, Singapore.

## Abstract

Age-related macular degeneration (AMD) is a leading cause of irreversible vision loss in ageing populations, with oxidative stress recognised as a key pathogenic driver. The dietary antioxidant and cytoprotectant, L-ergothioneine (ET), is avidly accumulated in many tissues, especially the eye. However its relationship to AMD has not been investigated.

Here, we examined ET’s distribution in ocular tissue and assessed circulating and intraocular ET levels in patients with neovascular AMD. Compared with ocularly-normal age-matched individuals, AMD patients exhibited significantly lower serum ET; elevated levels of ET metabolites, hercynine and ETSO₃, which may be generated by oxidative stress; and elevated levels of serum allantoin, a product of oxidative damage to urate in humans. Levels of ET in aqueous humour in AMD patients were marginally lower than cataractous patients who are already known to have significantly lower ET levels than healthy eyes. High ET levels were seen in human ocular tissues concentrating in regions vulnerable to oxidative injury, including the lens, retina, retinal pigment epithelium, and choroid, supporting a physiological protective role of ET in the eye.

These findings identify the strong association between low ET levels and AMD, warranting further studies to determine whether ET supplementation can modify AMD risk or progression.

## 1. Introduction

Rapidly aging populations across the globe, have fueled the rise in incidence of age-related diseases. Age is one of the greatest risk factors for many major diseases, including neurodegenerative and cardiometabolic diseases, many cancers and especially ocular disorders such as age-related macular degeneration (AMD), and an important contributory factor to these age-related diseases is the progressive accumulation of oxidative damage and mitochondrial dysfunction ^1–6^.

The eye is inherently vulnerable due to its high metabolic activity (especially the retina) and constant exposure to atmospheric oxygen levels, necessitating high levels of endogenous and dietary antioxidants as countermeasures ^2^. Exposure to environmental toxins and light, poor diet, or loss of metabolic homeostasis, can result in decreased levels of antioxidant defense and an increased generation of reactive oxygen species (ROS) in the eye, leading to damage to ocular tissues ^1,2,6–9^. Indeed, oxidative damage has long been proposed as a risk factor in the pathogenesis of AMD ^4^.

AMD is a leading cause of irreversible blindness in the elderly, accounting for 9% of people with blindness, a figure that will rise as aging populations increase ^6,7^. AMD is characterized by degeneration of the macula, resulting in blurry central vision and eventual vision loss, and in the advanced stage is classified into neovascular and non-neovascular (geographic atrophy; GA) forms. Current treatments for AMD are aimed at managing the late-stage complications of advanced disease, using intravitreal anti-VEGF injections for neovascular AMD, and anti-complement injections for GA. These therapies come with significant cost and treatment burden. Findings from the Age-Related Eye Disease Study (AREDS) and AREDS2 trials involving a formulation of nutritional and antioxidant supplements taken orally, were found to reduce the risk of progression from intermediate to advanced AMD ^8,9^. This suggests that there is potential for other antioxidants to reduce the risk of progression of AMD in the earlier stages of disease.

Ergothioneine (ET) is a dietary thiol/thione that cannot be made in the human body, but is avidly taken up and accumulated in all body tissues by a highly specific transporter OCTN1 ^10,11^. The high stability of ET and high retention (owing to renal reabsorption) in the body, facilitate strong tissue accumulation and suggest that ET may play an important physiological role ^12^. Supporting this, a wide range of studies has shown that ET possesses a wide range of cytoprotective properties including (but not limited to) antioxidant and anti-inflammatory roles, preserving mitochondrial function, and activating endogenous defense pathways (e.g. Nrf2, SIRTs) in a range of animal and cellular models of neurodegenerative and cardiovascular diseases and of aging, amongst others ^12–14^. Moreover, human studies have shown that lower plasma levels of ET are strongly correlated with a range of age-related diseases across different systems, including cognitive impairment and dementia ^15–17^, Parkinson’s disease ^13,18^, cardiovascular disease ^19^, and frailty ^20,21^.

Earlier studies have demonstrated that high levels of ET are present in the eyes of different animals, suggesting it may have a conserved role in protecting ocular tissue ^22,23^. Indeed, a prior study has found that progressively lower levels of ET in human eye lenses were associated with increasing severity of cataracts ^24,25^. As such, we hypothesize that ET levels may also play a role in the pathophysiology of AMD, another major age-related ocular disease. However, no study has examined the relationship between ET and AMD nor definitively examined the distribution of ET and its metabolites in the human eye using modern analytical methods. Such data may provide insights into the potential protective role of ET in prevention of ocular disorders. The present study examines the levels of ET in serum and aqueous humour (AH), via liquid chromatography-mass spectrometry, in AMD patients relative to controls and further characterizes the distribution of ET within ocular tissue by examining how ET differentially concentrates in the various ocular compartments in dissected post-mortem eye globes.

## 2. Materials & Methods

### Ethical approvals

Ethical approval for this study was obtained from the Institutional Review Board of Singapore Health Services, Singapore (protocol number: 2021-2058), and the study was conducted in compliance with the tenets of the Declaration of Helsinki.

### Chemicals and materials

ET-d_9_, hercynine, hercynine-d_9_, ET-sulphonate, and S-methyl ET standards were kindly provided by ERGOLD (Montreuil, France). ET (>99.9%) was kindly provided by Gene III (Nanjing, China). HPLC-grade acetonitrile and methanol were purchased from Fisher Scientific (MA, USA). All other chemicals were purchased from Sigma-Aldrich (MO, USA), unless otherwise stated.

### Cross-sectional study from patient samples

#### Serum samples (cohort I)

Serum samples were obtained from a research cohort of 200 deidentified participants from the Singapore Eye Research Institute (SERI). This included 100 participants with neovascular AMD (either typical neovascular AMD or polypoidal choroidal vasculopathy) from the Asian AMD Phenotyping Study, and 100 age-matched healthy controls without AMD from the Singapore Epidemiology of Eye Diseases (SEED) study ^26^. Participants with neovascular AMD had their diagnosis confirmed by clinical examination, and multimodal imaging with optical coherence tomography (OCT), fundus fluorescein angiography (FFA), and indocyanine green angiography (ICGA), and were treatment-naïve at the time of serum sampling. Venous blood was collected in clot activator tubes and left to sit at room temperature for 1h before subsequently centrifuging at 2000*g* for 10 min to obtain serum, which was stored at −80℃ until analysis of ET and related metabolites (ET and its metabolites are stable in human body fluids with storage at −80℃). Informed consent was obtained from all participants, and the ethics approval was obtained from the relevant institutional review boards (protocol numbers: 2009/788/A, R697/47/2009, R498/47/2006).

#### Aqueous humour (AH) samples (cohort II)

AH samples were obtained from a separate cohort of deidentified patients from Jichi Medical University, Shimotsuke, Japan. AH were obtained from 98 patients with treatment-naïve neovascular AMD. Diagnosis of neovascular AMD was confirmed by clinical examination and multimodal imaging with OCT, FFA, and ICGA. AH sampling in these eyes was performed in outpatient clinics, just before they received their first intravitreal anti-VEGF injections. AH samples were also obtained from 43 non-AMD control eyes at the time of cataract surgery. Samples were all aliquoted and stored at −80℃ until analysis. Informed consent was obtained from all participants, and the ethics approval was obtained from the Institutional Review Board of Jichi Medical University (JICHI20-127).

### Human Ocular Dissections

Healthy cadaveric eye globes (n=10) were obtained from the Lions Eye Institute for Transplant & Research (Tampa, FL, USA), and Saving Sight (Kansas City, MO, USA) eye banks (whole globes procured less than 24h after death, no AMD or other eye disease, no donors with dementia, Parkinson’s disease, or other neurodegenerative disease). The AH was aspirated from the anterior chamber and stored at −80℃. The cadaveric globes tended to be soft and hypotonous, and in some cases only small amounts of AH could be aspirated. After aspiration of AH, saline was injected into the anterior chamber to firm up the globe, to facilitate surgical dissection. The cornea, iris, lens (in phakic eyes), optic nerve, retina, retinal pigment epithelium (RPE), choroid and sclera were then dissected. Vitreous humour samples were obtained following dissection of the anterior segment. Due to difficulties in separating the RPE from the choroid, analyses were performed on the combined RPE-choroid tissue. All tissue samples were stored at −80℃ until analysis.

### Sample Preparation and Analysis by Liquid Chromatography-Mass Spectrometry (LC-MS/MS)

Serum, AH, and vitreous humour samples were prepared for LC-MS/MS analysis by adding 20μL of sample to 80μL of ice-cold methanol containing heavy-labelled internal standards (2μL of 25μM ET-d_9_, 1μL of 25μM hercynine-d_9_, and 1μL of 50μM ^13^C_1_^15^N_1_-allantoin). Samples were vortexed and incubated overnight at −20℃.

Cornea, iris, lens, optic nerve, retina, RPE-choroid, and sclera tissues from dissected postmortem globes were accurately weighed and transferred into Precellys tubes containing 3x zirconium oxide beads (2.8mm). 500μL of 4:1 methanol: ultrapure water (Arium Pro, Sartorius, Gottingen, Germany), containing heavy-labelled internal standards (as above) were added to each sample. The samples were then homogenized using the Precellys 24 Homogeniser (Bertin Technologies SAS, Montigny-le-Bretonneux, France) at 5000rpm for 3 cycles of 2 min, and repeated 6 times to ensure complete homogenization of all tissues. Samples were then incubated overnight at −20℃.

After overnight incubation, samples were vortexed and centrifuged at 14,000*g* (at 4℃) for 15 min. For dissected tissues, the Precellys zirconium oxide beads were removed prior to centrifugation. The supernatants were transferred to glass vials for drying under a stream of N_2_ gas at 40℃. Samples were reconstituted in 100μL of 90% methanol then diluted 10x with ultrapure water and transferred into silanized vial inserts for measurement by LC-MS/MS, which was carried out using an Agilent 1290 UPLC System coupled to an Agilent 6460 QQQ electrospray ionization (ESI) mass spectrometer (Agilent Technologies, CA, USA). Sample vials were maintained at 10℃ on the autosampler until analysis.

Analysis was performed according to ^27^ with minor modifications. Briefly, 2uL of extracted samples were injected onto a Cogent Diamond Hydride 2.0 column (Microsolv, NC, USA). Chromatographic separation was achieved using a gradient elution at 0.5mL/min for 15 min. Solvent A was acetonitrile containing 0.1% formic acid and Solvent B was ultrapure water containing 0.1% formic acid. The gradient elution commenced with 90% Solvent A: 10% Solvent B for 2 min, followed by a gradual adjustment to 40% Solvent A: 60% Solvent B over 8 min, and finally 100% Solvent B for 2 min.

### Ion Transitions and Mass spectrometer Parameters

Precursor to product ion transitions and fragmentor voltages (V)/collision energies (eV) for each compound were as follows: ET: 230.1 → 186, 103 V/9 eV; hercynine: 198.1 → 95.1, 94 V/21 eV; ETSO_3_: 278 → 154, 120 V/15 eV; S-methyl-ET: 244.1 → 141, 92 V/17 eV, allantoin: 159 → 116, 70 V/2 eV; ET-d9: 239.1 → 195.1, 98 V/9 eV; hercynine-d9: 207.2 → 95.1, 97 V/21 eV; and allantoin-4-^13^C,1-^15^N: 161 → 118, 70 V/2 eV.

### Statistical Analysis

Statistical analyses were performed using GraphPad Prism (version 10). Data are expressed as mean ± standard error of the mean. For study cohort I, Pearson’s correlation coefficient was used to calculate the association between age and ET levels. The Mann Whitney U test was used to establish significance, where *p* < 0.05 is considered statistically significant.

## 3. Results

### 3.1. Population Demographics

Patient samples using in this study were obtained from two separate cohorts. Serum samples were analyzed from cohort I, which consisted of patients with neovascular AMD and healthy age-matched controls (with n=100 per group). AH samples were analyzed from cohort II, which similarly consisted of patients with neovascular AMD (n=98) and controls without AMD but undergoing cataract surgery (n=43). Demographic data of both study cohorts are provided in Table 1. No significant differences were observed in age and gender between the control and AMD groups in either cohort.

**Table 1.** Demographic Data of Study Cohorts I (serum samples; Singapore) & II (AH samples; Japan).

| Baseline Demographics | Control Group | AMD Group |
| --- | --- | --- |
| *Serum analysis cohort* |  |  |
|  | (n = 100) | (n = 100) |
| Age (years) | 70.8 ± 7.9 | 72.3 ± 9.3 |
| Gender |  |  |
| - Male, n (%) | 61 (61.0) | 57 (57.0) |
| - Female, n (%) | 39 (39.0) | 43 (43.0) |
| Ethnicity |  |  |
| - Chinese, n (%) | 89 (89.0) | 88 (88.0) |
| - Malay, n (%) | 0 (0.0) | 9 (9.0) |
| - Indian, n (%) | 11 (11.0) | 1 (1.0) |
| - Others, n (%) | 0 (0.0) | 2 (2.0) |
| *Aqueous humour analysis cohort* |  |  |
|  | (n = 43) | (n = 98) |
| Age (years) | 73.65 ± 8.13 | 73.98 ± 8.80 |
| - 80+, n (%) | 13 (30.2) | 27 (27.6) |
| - 65 – 79, n (%) | 21 (48.8) | 58 (59.2) |
| - 50 – 64, n (%) | 9 (21.0) | 13 (13.2) |
| Gender |  |  |
| - Male, n (%) | 31 (72.1) | 75 (76.5) |
| - Female, n (%) | 12 (27.9) | 23 (23.5) |

### 3.2. Lower ET levels were observed in AMD patients

We first assessed the association between age and serum ET levels in healthy controls and AMD patients independently. Similar to prior studies ^15^, significant inverse correlations between blood ET levels and age were observed in both healthy controls (r = −0.253; *p* = 0.0335; Fig. 1A) and AMD patients (r = −0.203; *p* = 0.0455; Fig.1B). ET levels in healthy controls were not associated with gender, or incidence of hypertension or hypercholesterolaemia (data not shown).

**Figure 1:**
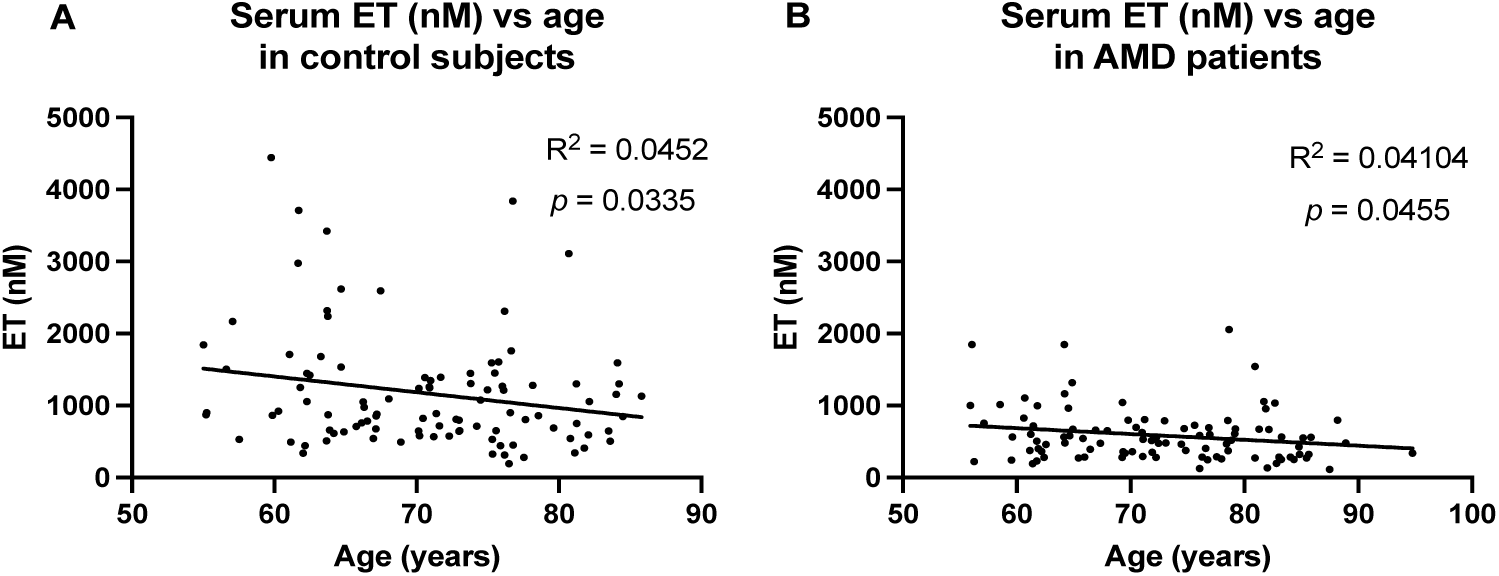
A significant (p = 0.0335 & 0.044) inverse correlation was observed between serum ET levels and age of individuals in the **(A)** healthy controls (n=100) and **(B)** AMD patients (n=100).

Serum levels of ET and its metabolites hercynine, and ET sulphonate (ETSO_3_), were assessed using LC-MS/MS and levels were compared between healthy controls and AMD patients. Although the AMD group was comprised of polypoidal choroidal vasculopathy (PCV) and typical neovascular AMD patient subtypes, there was no significant difference observed between levels of any marker between these groups hence for the purpose of this study these are combined (*Suppl. Fig. S1*). However, levels of ET in serum of AMD patients were observed to be significantly lower (617.4 ± 50.58 nM for ET) than age-matched healthy controls (1169 ± 80.88 nM; Fig. 2A; *p < 0.0001*). Interestingly, significantly higher serum levels of hercynine and ETSO_3_ were observed in the AMD group (40.86±2.79 nM; *p < 0.001* and 315.7±65.79 nM; *p < 0.0001*, respectively) as compared to the healthy controls (35.88±4.27 nM and 59.13±18.3 nM, respectively; Fig. 2B and 2C).

**Figure 2:**
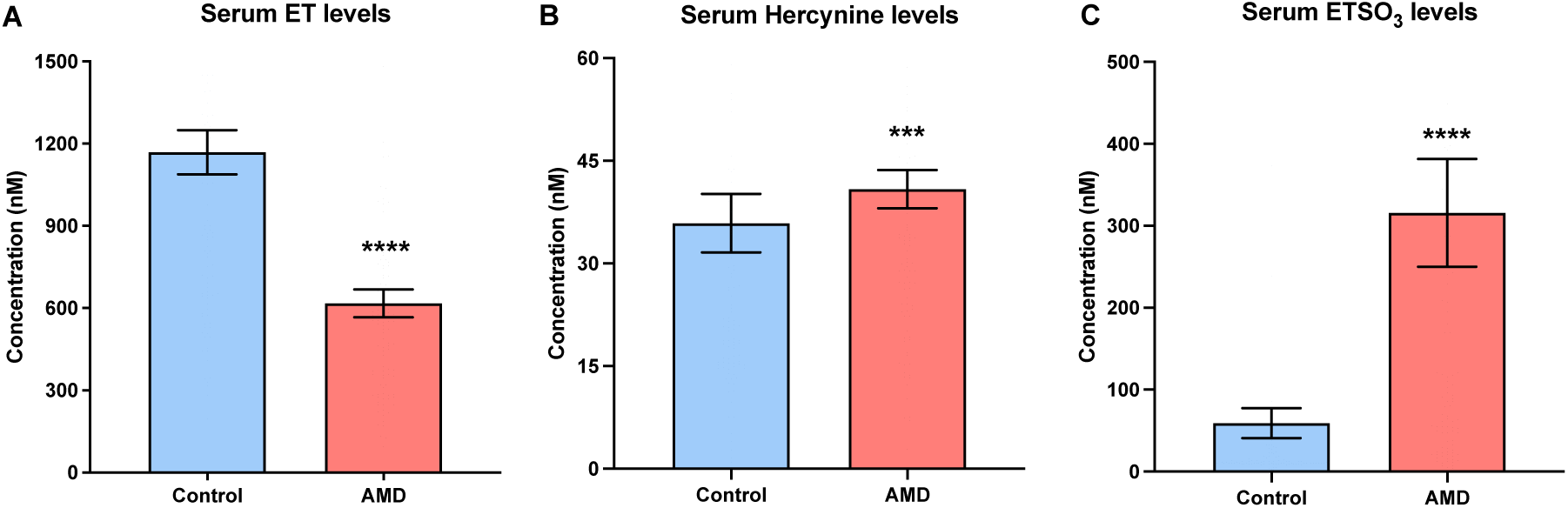
Comparison of mean (± SEM) serum levels of **(A**) ET, and its metabolites **(B)** hercynine, and **(C)** ETSO_3_ between normal controls (n=100), AMD patients (n=100). Serum levels of ET were significantly lower, while serum hercynine and ETSO_3_ were significantly higher in AMD patients versus healthy controls (Mann-Whitney U test; ***p<0.001, and ****p < 0.0001).

We also examined allantoin levels as an established biomarker of oxidative damage; allantoin is produced when oxygen radicals and other reactive oxygen species attack uric acid ^2^. Serum allantoin levels were also significantly elevated in the AMD group; 37.08±2.12 nM; *p < 0.0001* compared with healthy controls; 4.83±0.17 nM; Fig. 3. No significant differences were observed between AMD subtypes (*Suppl. Fig. S1*).

**Figure 3:**
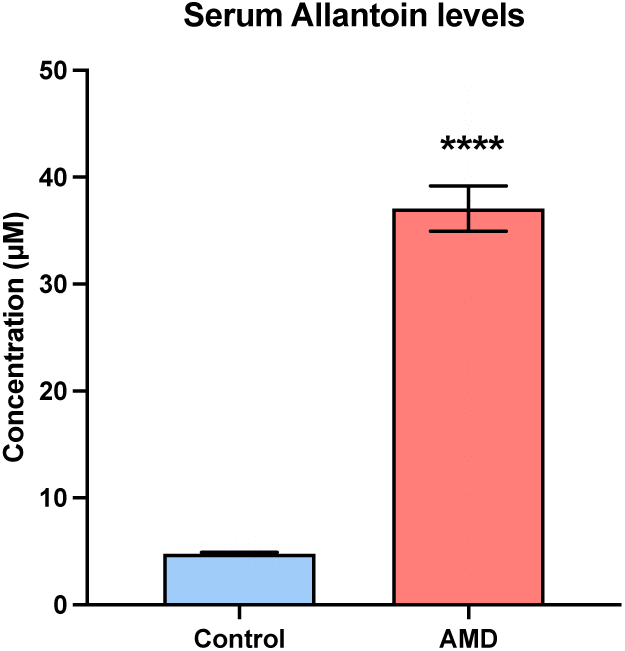
Comparison of mean (± SEM) serum levels of allantoin between normal controls (n=100), AMD patients (n=100). Serum levels of allantoin were significantly higher in AMD patients versus healthy controls (Mann-Whitney U test; ****p < 0.0001).

### 3.3. Aqueous humor levels of ET and metabolites in cohort II

Given the significant differences in serum ET levels between healthy controls and AMD patients, we sought to examine if these systemic differences could also be observed in AH within the eye. Collection of AH samples is an invasive procedure. For the neovascular AMD patients, AH sampling from the anterior chamber was performed just before anti-VEGF injection, as part of the same overall procedure. For non-AMD control participants, it was not deemed feasible to sample AH, unless they were already undergoing an intraocular procedure. Therefore, we selected non-AMD control participants who were undergoing cataract surgery, and AH sampling was performed at the start of the procedure. Hence, non-AMD controls in this study were defined as patients without macular abnormalities but with cataracts.

In the AH, a decreasing trend of ET was observed in AMD patients relative to cataractous patients (Fig. 4A), however, these differences were not statistically significant. Interestingly, an increase in ETSO_3_ and hercynine levels were observed in the AH of AMD patients, however this was only significantly higher for hercynine (*p*<0.05; Mann-Whitney; Fig. 4B-C). No changes were observed in allantoin levels in the eye between all groups (Fig. 4D).

**Figure 4:**
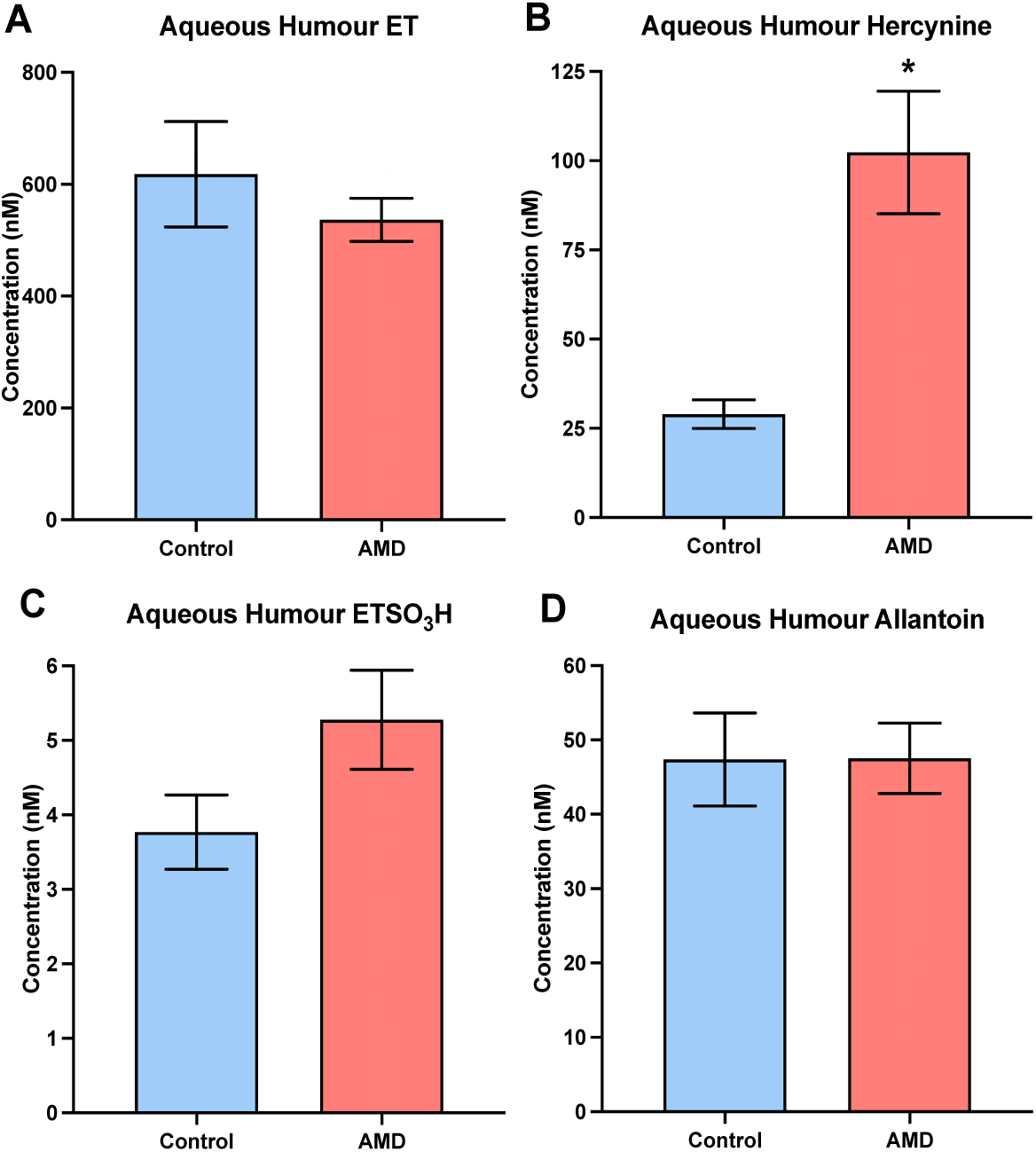
Comparison of the mean (± SEM) levels of **(A)** ET, **(B)** hercynine, **(C)** ETSO_3_, and **(D)** allantoin in AH between normal controls (n=42) and AMD patients (n=98; comprising PCV/tAMD). Despite a decreasing trend of ET and increasing trend of ETSO_3_ in the AH of AMD patients (vs. controls), this was not significant. Hercynine levels in AMD patients however, were found to be significantly elevated versus the control group (Mann-Whitney U test *p<0.05).

### 3.4. Differential accumulation of ET and its metabolites in various ocular tissues

Prior studies have shown that animal eyes accumulate high levels of ET ^22,23^. However, no studies to date have examined ET accumulation in human eyes (with the exception of human tear fluid and AH ^28^), nor its differential uptake in the various tissues/regions of the eye. To examine this we extracted and quantified ET and its metabolites in different ocular tissues of healthy (no AMD or other eye diseases) human eye globes.

ET was detectable across all compartments of the human eye that were analysed (Fig. 5; *Suppl. Fig. S2*) but highly concentrated in certain regions. The lens had by far the highest levels of ET at 408.70±126 ng ET /mg tissue (wet weight), approximately fifty times higher than the RPE and choroid, which was the second highest level at 12.47 ± 2.98 ng ET/ mg tissue (Fig 5A). This is followed by the, cornea (5.45 ± 1.30 ng ET/ mg tissue), retina (4.95 ± 1.08 ET ng/ mg tissue), and iris (1.20 ± 0.17 ng ET/ mg tissue). Similar trends were seen with ET metabolites, with the lens having the highest concentrations of hercynine and ETSO_3_ (Fig. 5B-C) followed by RPE & choroid, cornea, and retina. On the other hand, the highest levels of allantoin were observed in the RPE & choroid regions with lower levels observed in the lens, cornea, and retina (Fig. 5D). Another ET metabolite, S-methyl ergothioneine, was also analysed however for most regions of the eye this was below the limits of quantification. We also analysed ophthalmic acid (OPA), which is a suggested indicator of glutathione utilisation ^29^, and the levels were closely correlated to ET (*Suppl. Fig. S3*).

**Figure 5:**
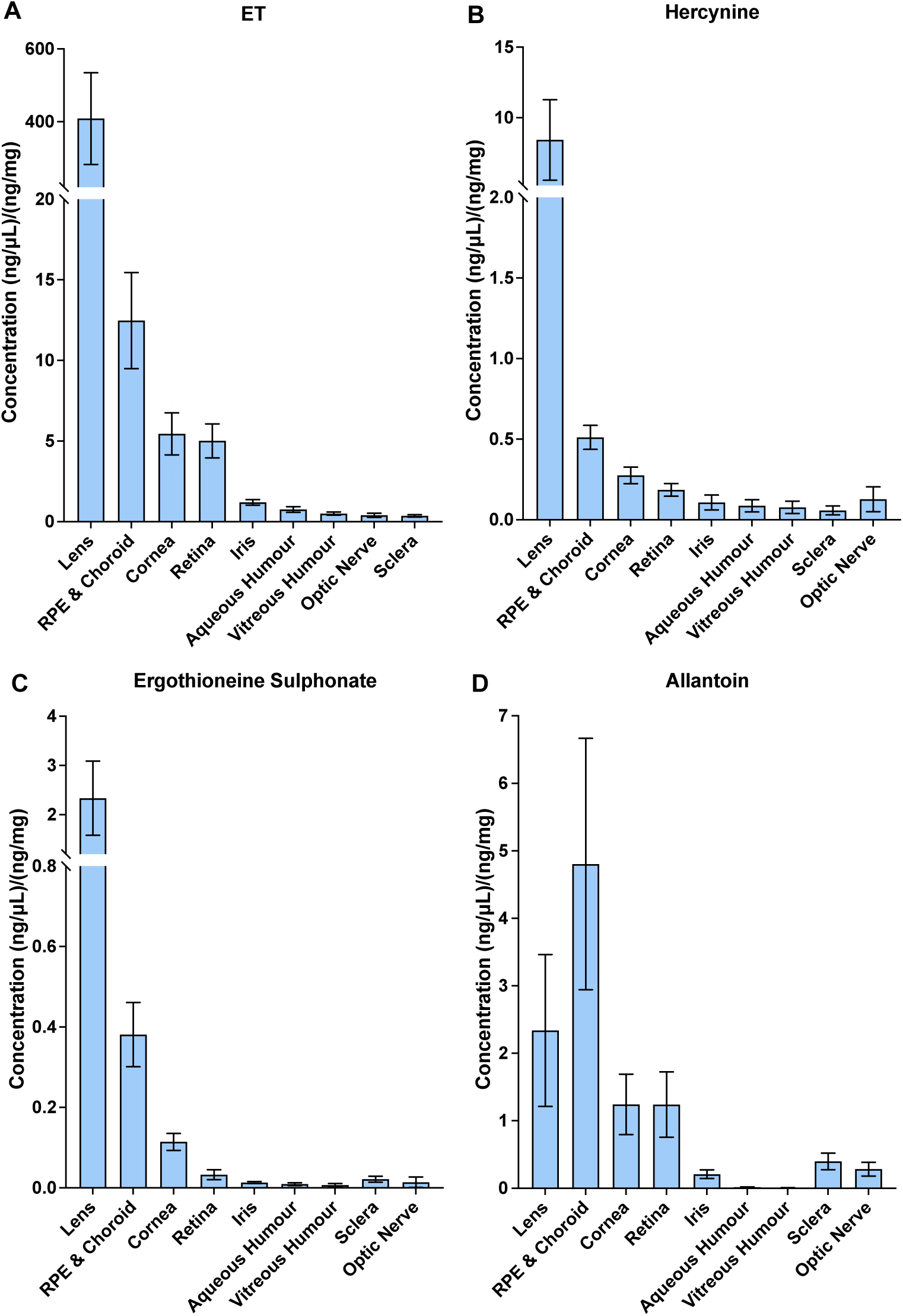
Levels of **(A)** ET, **(B)** hercynine, **(C)** ergothioneine sulphonate (ETSO_3_), and **(D)** allantoin within different ocular tissues/regions of the human eye. All data points are expressed as amount of ET detected amount (ng) per mg wet weight of tissue or µL liquid. For ET and metabolites, the highest levels were found in lens, and RPE and choroid.

## 4. Discussion

AMD is one of the leading causes of irreversible blindness in elderly individuals, affecting one in eight people over the age of 60 and around 200 million individuals worldwide ^6^. With rapidly aging populations across the globe, this age-related disorder is projected to rise further to around 300 million by 2040 ^7^. There is no curative treatment for this progressive degeneration, but existing treatments are aimed at treating its late stage complications, and preventing vision loss ^30^. Prior studies have shown that the dietary antioxidant and physiological cytoprotectant, ET, is able to accumulate at high levels in the eye of animals ^23^, however to date no studies have evaluated the relation between this compound and AMD.

As lower plasma levels of ET have been previously established to be associated with increased incidence of a range of age-related disorders, including cognitive impairment and dementia, Parkinson’s disease, cardiovascular disease and frailty ^13^, we assessed if a similar association could be identified between blood ET levels and AMD. Indeed, lower serum ET levels were observed in patients with neovascular AMD in this study (no difference was observed in ET levels between different AMD subtypes; *Suppl. Fig. S1*). It should be noted that we previously established that there is no difference between plasma and serum levels of ET (drawn at identical time points in another cohort), suggesting that ET levels are not impacted by the complement system (*Suppl. Fig. S4*).

The retina is one of the tissues with the highest oxygen consumptions in the human body ^2^, hence it is not surprising that oxidative stress plays a key role in the pathophysiological progression of AMD ^4,6,8,31^. In addition to decreased levels of ET in AMD patients, elevated levels of allantoin and the ET metabolites, hercynine and ETSO_3_, were observed. The elevation of allantoin, an established biomarker of oxidative damage, produced by oxidative modification of uric acid ^2,32,33^, in AMD patients suggests that these patients were exposed to a higher level of oxidative stress. This is supported by higher levels of hercynine and ETSO_3_, ET metabolites, which have been suggested to arise through oxidative breakdown of ET ^34,35^. Typically, blood levels of ETSO_3_ in healthy individuals are relatively low or undetectable by our LC-MS/MS methods, however the levels are significantly elevated in AMD patients. Hence, we postulate that AMD patients are exposed to higher levels of oxidative stress, and one of the factors leading to decreased serum ET may be a higher utilisation, hence the elevation in hercynine and ETSO_3_. However, we cannot pinpoint if this elevated oxidative stress is systemic or localised to the eye, since it is possible that these oxidative products originating from diseased tissues are then released systemically into circulation for excretion, especially due to the vascularity of the choroid.

The cross-sectional nature of this study does not establish if the lower ET levels precede or follow AMD onset. However, prior studies have shown that low blood ET levels precede and predict onset of other age-related disorders ^17,36^, and if ET levels in serum are lower prior to disease onset this would suggest that low blood (and hence tissue) levels of ET are a possible modifiable risk factor for the development of AMD, and possibly supplementation may help prevent onset or slow the progression of disease. However, further longitudinal and/or clinical studies are needed to validate this. It is also possible that lower serum ET levels may be due to other factors such as dietary changes, concomitant illness, or its utilisation (e.g. as an antioxidant) resulting from the disease. Regardless, this highly significant decrease in serum ET suggests an association between ET and AMD, especially since the eye is known to accumulate ET.

Serum levels may provide a general indication of ET levels in the body. However, it is important to assess if this association is reflected within the eye. AH may be collected prior to cataract surgery or prior to intravitreal anti-VEGF injections in AMD patients. The AH is known to be a reservoir of endogenous and dietary antioxidants (such as glutathione and glutathione peroxidase, catalase, superoxide dismutase, ascorbic acid, lutein, zeaxanthin ^37,38^) which help to protect the eye from oxidative damage. Despite a decreasing trend of ET levels in the AH of AMD patients, this was not significantly different from cataractous patients. However, a prior metabolomics study identified that cataractous patients had significantly lower levels of ET (as well as glutathione and ascorbate), relative to ocularly normal AH (post-mortem eyes) ^39^. Indeed, elevated levels of oxidative stress and decreased levels of ocular antioxidants are key pathological findings of cataracts ^40^. Significantly lower levels of ET have also been previously been reported in cataractous lenses, (although this study may be worth revisiting since this very old study harnessed old and possibly inaccurate analytical techniques) ^24,25^.

Due to the invasive nature of AH sampling which can only be performed alongside other clinically-indicated ocular procedures we acknowledge that our study has limitations. One is the lack of normal AH controls, hence we substituted a control with no macular abnormalities, but had clinically significant cataracts. Secondly, AMD is primarily a degenerative condition of the retina and RPE and the AH in the anterior chamber is compartmentalized from these posterior segment tissues; however, measurement of intracellular ET levels is not possible *in vivo*. The next closest sample would be the vitreous humour, however this is also far less accessible and even more invasive than AH sampling, and further limits options for control samples. Lastly, we also acknowledge the difference in population demographics between serum and AH samples (although they each have their own respective controls). Ideally matched serum and AH samples for the same AMD patient would better demonstrate if systemic differences in ET are matched intraocularly to AH levels. Regardless, the decreasing trend of ET levels in AH of AMD patients relative cataractous patients which are potentially already significantly lower in ET levels than normal, suggests that ET levels are significantly decreased in macular degeneration. Like in serum, elevated levels of hercynine and ETSO_3_ were also observed in AH of patients with AMD, suggesting that the eye is indeed under a state of elevated oxidative stress. Interestingly, unlike serum, the allantoin levels were not different between AMD and cataractous patients. Again the lack of difference in allantoin between these groups may be due to already elevated levels in cataracts, since prior studies have found that cataractous patients have elevated levels of uric acid in the AH ^41^ and cataracts are a condition with substantial oxidative stress; hence the cataractous samples are likely to have elevated levels of allantoin relative to healthy eyes. Another scenario may be that allantoin, is be rapidly cleared from the eye into circulation accounting for elevated serum levels, since its levels turn over rapidly in the human body^32^.

Due to the high vulnerability of the eye to oxidative stress, high levels of endogenous and dietary antioxidants are known to accumulate in ocular tissues such as glutathione in lens, and lutein and zeaxanthin in the retina ^42,43^. Prior studies have shown that ET was found to accumulate in bovine, porcine, mouse, and squirrel eyes ^22,23,44^, however no studies have yet examined the accumulation and distribution of ET in the human eye (although an earlier study examined metabolites in primate retina, confirming ET as the third most enriched metabolite measured in the macula ^45^, and our earlier study revealed that ET is present in human tear fluids ^28^). Studying the differential accumulation of ET in the various ocular segments may provide insight to the role ET plays in the eye. Indeed ET was present in all regions of the eye but especially high levels of ET could be found in certain regions of the eye that are exposed to high levels of oxidative stress, including the lens, retina, RPE, and choroid, which are tissues either exposed to atmospheric levels of oxygen or have very high energy demands. Trends observed for the metabolites of ET, hercynine and ETSO_3_, were found to closely follow that of ET, albeit at lower concentrations. The levels of allantoin in different eye compartments had a very different trend to ET and its metabolites, with the highest levels observed within the choroid.

Compared to AH from live patients, the levels of ET obtained in the AH and vitreous humour from the dissected cadaver globes were quite low. Indeed, metabolomic analyses of living and cadaveric eye samples have identified huge differences ^46^. For the vitreous humour samples, it is possible that some of the saline injected into the anterior chamber during cadaveric globe dissection went through the zonules into the posterior segment, and could have resulted in some dilution of the vitreous samples.

The avid accumulation of ET in the eye suggests that, alongside other antioxidants, ET may play a role in alleviating the ocular oxidative burden. Decreased levels of ET in the body and eye may be a risk factor for ocular disorders including AMD and cataracts. These findings also suggest that ET administration could be beneficial in preventing or possibly treating ocular disorders such as AMD. Contrary to many other antioxidants, our earlier studies in a mouse model demonstrated that orally administered ET is not only avidly accumulated by the body but raises levels in the eye after just 7 days of feeding ^23^. A recent study screening a library of antioxidants to protect RPE against oxidative stress, identified that ET was one of the most potent cytoprotective agents, capable of maintaining retinal structure in mice under oxidative stress through activation of the Nrf2 pathway ^14^. In addition to its antioxidant and anti-inflammatory properties, another key mechanism by which ET may protect the eye is through preservation of mitochondrial function. Mitochondrial dysfunction, particularly in the RPE, is a key hallmark of AMD, and is associated with increased ROS, mitochondrial DNA damage, protein aggregation and inflammation ^47,48^. Intracellularly, ET is taken up and accumulated in the mitochondria ^49^, and has been shown to prevent damage to mitochondrial DNA ^50^ and mitochondrial dysfunction in cell and animal models ^51,52^. ET has also been shown to protect the vascular system ^53,54^ and is associated with decreased risk of cardiovascular diseases ^19^, which are a major risk factor for AMD, and may also be relevant for PCV, a variant of neovascular AMD, with common underpinnings of vascular dysfunction (hence the application of anti-VEGF therapies). There have also been suggestions that co-application of antioxidants with anti-VEGF (including phototherapies, siRNAs, or drugs) may improve the effectiveness of these treatments ^55^, and ET is worth considering in this regard.

Further studies are needed to elucidate whether ET is indeed able to prevent or treat AMD and other ocular disorders and determine the mechanisms by which it may act. However, the current study demonstrates the association of lower ET levels with AMD and supports further investigation to evaluate whether ET may play a protective role in preventing or treating AMD or age-related ocular disorders.

## 5. Conclusions

High levels of ET are known to accumulate in the eye and this reflects the high antioxidant demand of metabolically active and oxidatively stressed ocular tissues, particularly the RPE-choroid complex. The present study identifies a strong association of lower serum (and likely AH) ET levels with AMD, suggesting that lower ET levels may be a risk factor for, and/or accelerate the pathological progression of AMD. Correlation does not, of course, confirm causation and further studies are needed to establish any preventative and therapeutic potential of ET (which is safe for human consumption; designated as GRAS [Generally Recognised As Safe] by U.S. Food and Drug Administration) to counteract AMD and other ocular disorders, which are an increasing problem in our aging societies.

## Acknowledgements

The authors wish to thank ERGOLD (Montreuil, France) for provision of L-ergothioneine, hercynine, and ETSO_3_ analytical standards used in these studies and Gene III (Nanjing, China) for L-ergothioneine, as well as Huifen Lin and Gary SL Peh, Singapore Eye Research Institute, for their assistance with cadaveric eye globe tissues.

## Author contributions

I.K.C., Z.F., R.M.Y.T., L.Z., T.E.T., B.H., K.Y.C.T., C.M.G.C. conceived and designed research studies; L.Z., Y.Y., C.Y.C., T.E.T., C.M.G.C. provided/obtain human samples for analysis; I.K.C., Z.F., X.L., R.M.Y.T. conducted experiments/analyses; I.K.C., Z.F. L.C., R.T.M.Y., B.H. analysed and tabulated data; I.K.C., Z.F., R.T.M.Y., B.H., X.S., K.Y.C.T. C.M.G.C., T.E.T. interpreted results; I.K.C., Z.F., L.C. prepared figures; Z.F., I.K.C., L.C., R.T.M.Y. drafted the manuscript; I.K.C., B.H., T.E.T., C.M.G.C., X.S.; provide manuscript revisions and feedback. I.K.C., R.M.Y.T., B.H., T.E.T., L.Z.; sourced for funding to conduct the studies. All authors read and approved the final manuscript.

## Data availability statement

All data generated from this study are including the article, supplementary figures or available on request from the corresponding authors.

