## Supplementary material for "Are low ergothioneine levels a risk factor for age-related macular degeneration and other ocular disorders?": Suppl. Fig.

**Supplementary figures:**

***Figure S1:*** *No significant differences were observed between sub-variants of AMD (PCV and tAMD) in ET, ET-related metabolites and allantoin (ns: not significant; Mann-Whitney U test).*

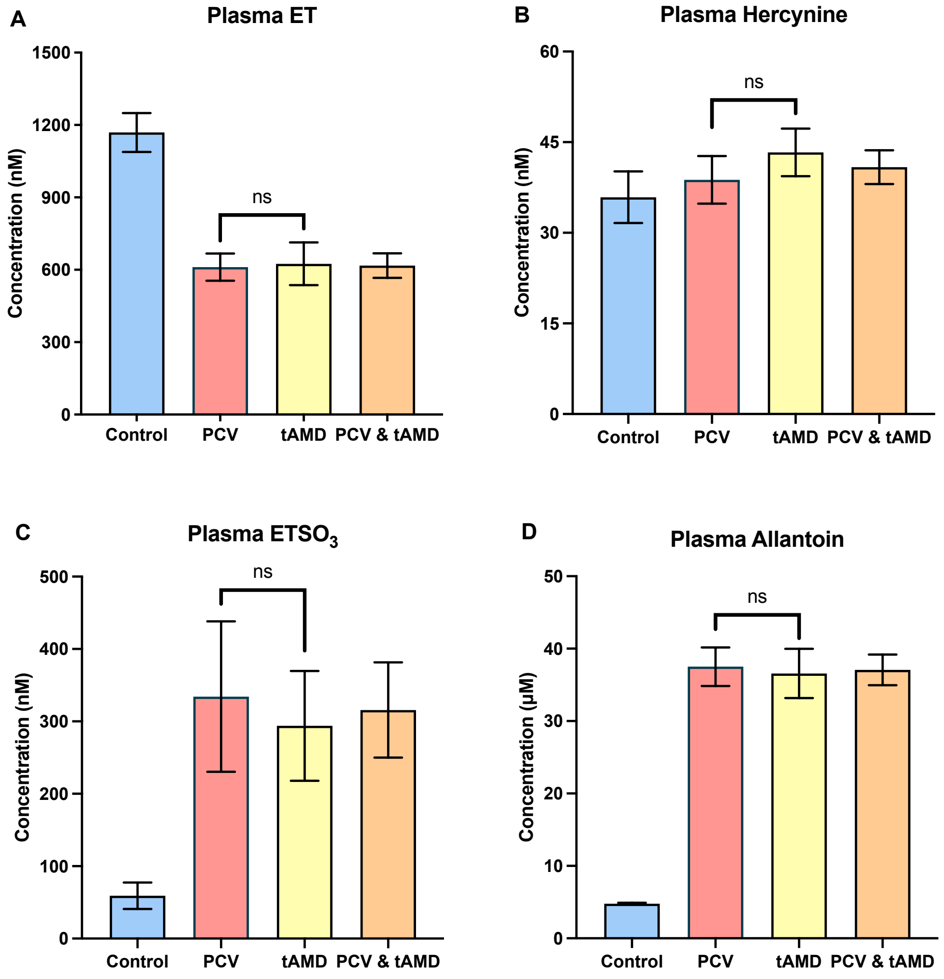

***Figure S2.*** *Tissue levels of ET, hercynine, $ETSO_3$ and allantoin levels in various ocular compartments. The RPE and choroid combined comprised samples that could not be separated during the dissection.*

| Compartments | ET | Hercynine | $ETSO_3H$ | Allantoin |
|---|---|---|---|---|
| | **(ng/mg)** | **(ng/mg)** | **(ng/mg)** | **(ng/mg)** |
| *Lens* | *408.7 ± 126* | *9.8 ± 3.3* | *1.9 ± 0.6* | *3.4 ± 1.6* |
| | **(pg/mg)** | **(pg/mg)** | **(pg/mg)** | **(pg/mg)** |
| *RPE & Choroid* | *12210 ± 3.9* | *375.4 ± 107.5* | *249.4 ± 66.4* | *6026 ± 2220* |
| *Choroid* | *5817 ± 1365* | *906 ± 261.5* | *354.1 ± 131.5* | *24240 ± 9461* |
| *Cornea* | *5384 ± 1328* | *265.4 ± 66.9* | *89.2 ± 19.7* | *1804 ± 648.7* |
| *Retina* | *4951 ± 1084* | *165.3 ± 50357* | *25.7 ± 10.6* | *1799 ± 701.1* |
| *RPE* | *2747 ± 962.8* | *512.1 ± 237.4* | *122.4 ± 66.8* | *8471 ± 3166* |
| *Iris* | *1099 ± 182.8* | *45.1 ± 5.9* | *8.5 ± 1.8* | *306.5 ± 93.1* |
| *Aqueous* | *754.4 ± 153.6* | *24.2 ± 4.8* | *3.3 ± 3.3* | *229.6 ± 130.2* |
| *Vitreous* | *428.6 ± 74.6* | *15.9 ± 3.5* | *4.2 ± 4.2* | *200.3 ± 114.5* |
| *Sclera* | *312.6 ± 71.2* | *27.4 ± 7.3* | *14.6 ± 6.9* | *580.1 ± 176.7* |
| *Optic Nerve* | *223.6 ± 39.6* | *15.4 ± 1.8* | *4.9 ± 4.7* | *415.4 ± 147.2* |

***Figure S3:*** *Levels of S-methyl ergothioneine (SMET; left) and ophthalmic acid (OPA; right) in different compartments of the eye. SMET levels were typically close to limits of detection. OPA is a tripeptide suggested to be involved in glutathione regulation whose distribution levels follow closely to that of ET.*

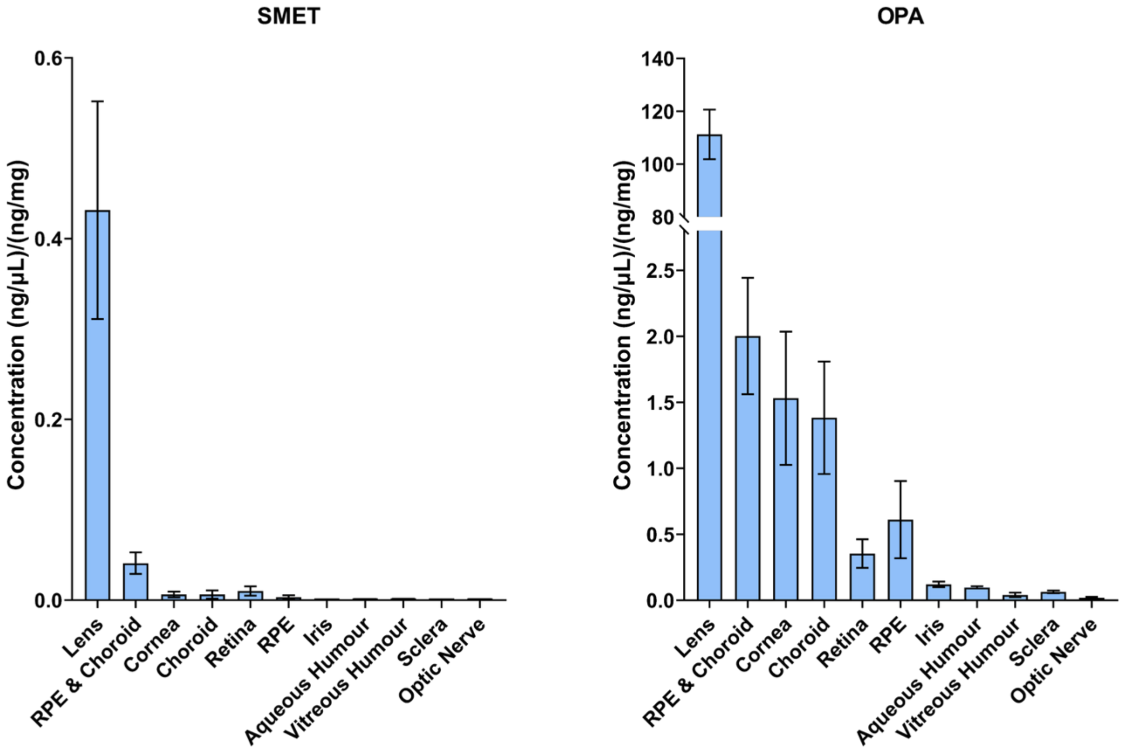

***Figure S4:*** *Correlation of plasma and serum levels of ET. We have shown that serum levels of ET are very highly correlated to plasma ET levels (collected consecutively from the same subject)* [27].

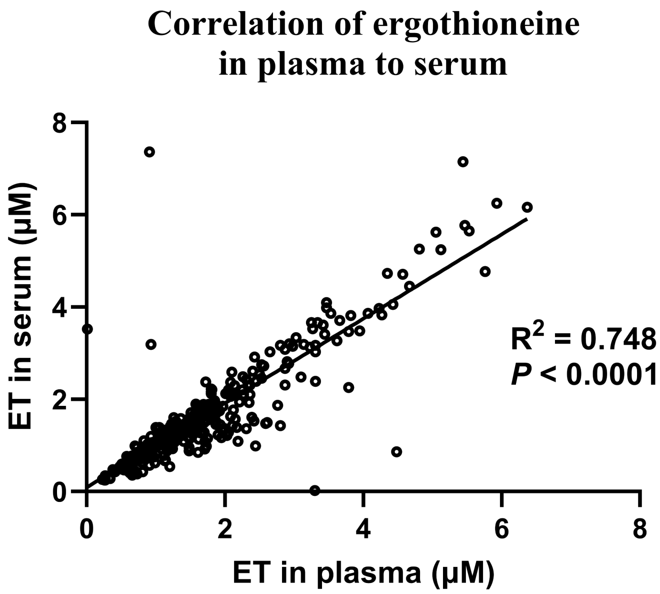
